# Association between the hemoglobin albumin lymphocyte and platelet score and chronic kidney disease: insights from patient data and animal models

**DOI:** 10.64898/2026.06.20.26356118

**Authors:** Wenyu Zhang, Yashi Wang, Wenfeng Ye, Yiman Wang, Xuejing Chen, Bingwu Zhao, Xueqin Zhang, Zhiqiang Chen

**Affiliations:** Hebei University of Chinese Medicine, Shijiazhuang 050091, Hebei, China; The First Affiliated Hospital of Hebei University of Chinese Medicine, Shijiazhuang 050200, Hebei, China

## Abstract

**Introduction:** The hemoglobin, albumin, lymphocytes and platelets (HALP) score, a novel nutritional and inflammatory biomarker, has been used in various chronic disease studies. However, the relationship between the HALP score and chronic kidney disease (CKD) remains poorly elucidated. This study aimed to explore the possible association between the HALP score and CKD.

**Methods:** Our analysis encompassed 25,160 adult participants drawn from NHANES cycles spanning 2009 through 2018. Weighted multivariable logistic regression and generalized additive models (GAMs) were employed to evaluate the independent associations between the HALP score and CKD, albuminuria, and low-estimated glomerular filtration rate (eGFR). Threshold effects were examined using two-piecewise linear regression. Subgroup and sensitivity analyses were performed to assess robustness. Receiver operating characteristic (ROC) curve analyses were applied to compare the discriminative capacity of the HALP score with the prognostic nutritional index (PNI), systemic immune-inflammation index (SII), lymphocyte-to-monocyte ratio (LMR), and platelet-to-lymphocyte ratio (PLR). The clinical findings were further validated in a 5/6 nephrectomy rat model.

**Results:** After adjustment for multiple confounders, higher HALP scores were inversely associated with the risk of CKD (OR = 0.97, 95% CI: 0.94–0.99) and albuminuria (OR = 0.97, 95% CI: 0.93–0.99). However, after full adjustment for demographic characteristics, physical examination indices and laboratory parameters (Model 3), the correlation between the HALP score and low-eGFR was no longer statistically significant. Non-linear analyses revealed a threshold effect, with CKD risk declining as the HALP score increased up to an inflection point of 52.43 (OR = 0.97, 95% CI: 0.95–0.99), beyond which no further protective effect was observed. A similar threshold effect was identified for albuminuria. Subgroup and interaction analyses indicated no meaningful effect modification by age, sex, BMI, hypertension, or diabetes. Sensitivity analyses confirmed the robustness of the results. ROC analysis demonstrated that the HALP score showed superior discriminative ability for CKD and albuminuria compared with PNI, SII, LMR, and PLR. In the animal experiment, CKD model rats exhibited significantly lower HALP scores than controls. Inverse correlations were observed between the HALP score and serum creatinine (Scr), blood urea nitrogen (BUN), and urinary albumin-to-creatinine ratio (UACR), with UACR showing the strongest correlation, which was consistent with the clinical findings.

**Conclusion:** Lower HALP scores are independently associated with increased prevalence of CKD and albuminuria. As an affordable and readily measurable biomarker, the HALP score may facilitate CKD risk assessment.

## 1 Introduction

Chronic kidney disease (CKD) represents a critical public health concern globally, impacting several hundred million people and serving as a significant driver of disease burden and mortality worldwide. [1] A critical clinical challenge is that CKD is frequently asymptomatic in its early stages, which delays timely diagnosis. By the time clinical manifestations emerge, significant and often irreversible kidney injury has already occurred. [2] Therefore, early detection, prompt intervention, and effective management are essential for reducing the disease burden, highlighting the importance of identifying reliable risk markers for CKD progression. Accumulating evidence suggests that CKD progression is closely linked to persistent subclinical inflammation, a key mediator of kidney injury and renal function deterioration. [3] In addition to inflammation, malnutrition is also a recognized contributor to CKD development. [4] As the disease progresses, malnutrition tends to worsen, further adversely affecting patient outcomes. A bidirectional “inflammation-malnutrition” interaction has thus been proposed, suggesting that composite indicators integrating both dimensions may better capture CKD risk than single markers. [5] Indeed, growing evidence has demonstrated that several nutrition- and inflammation-related markers can serve as reliable indicators of CKD, including the systemic inflammation response index (SIRI) [6] and the advanced lung cancer inflammation index (ALI). [7] Initially formulated as a prognostic indicator for individuals with gastric malignancies, the HALP score simultaneously captures both the body’s inflammatory state and its nutritional condition concurrently. [8] Subsequently, its prognostic utility has been validated in a range of kidney disorders, such as diabetic nephropathy (DN), [9] immunoglobulin A nephropathy (IgAN), [10] lupus nephritis, [11] and for predicting clinical outcomes in dialysis patients. [12,13] However, existing research on the HALP score in renal disease has predominantly focused on specific subtypes such as DN and IgAN, as well as dialysis populations, while evidence from general-population cohorts remains scarce. In addition, experimental evidence verifying the biological pathways through which the HALP score relates to CKD remains limited. To address this gap, we utilized NHANES data to examine the associations between the HALP score and CKD, albuminuria, and low- eGFR. Our aim was to establish a reliable biomarker for CKD detection and management. Furthermore, a 5/6 nephrectomy rat model was established to validate the clinical findings.

## 2 Materials and methods

### 2.1 Data sources

Data analyzed in this study were obtained from NHANES, a continuous, nationally representative health survey administered by the NCHS that employs a complex multistage probability sampling framework. Ethical approval and written informed consent had already been secured under the original NHANES protocol; no further review was therefore required for this secondary analysis of de-identified public-use files.

### 2.2 Data collection and measures

Data from NHANES 2009–2018 were utilized for this analysis, including 49,693 participants. Exclusion criteria comprised participants younger than 20 years (n = 20,858), those who were pregnant (n = 315), and individuals with missing serum creatinine (Scr) or urinary albumin-to-creatinine ratio (ACR) data (n = 3,201). Subjects with insufficient information for HALP score computation (n = 83) were also omitted, resulting in 25,236 eligible participants. To address the potential confounding effects of renal replacement therapy, participants undergoing hemodialysis or peritoneal dialysis (n = 76) were further excluded. Consequently, the final analytic cohort included 25,160 participants (Fig 1).

**Fig 1.**
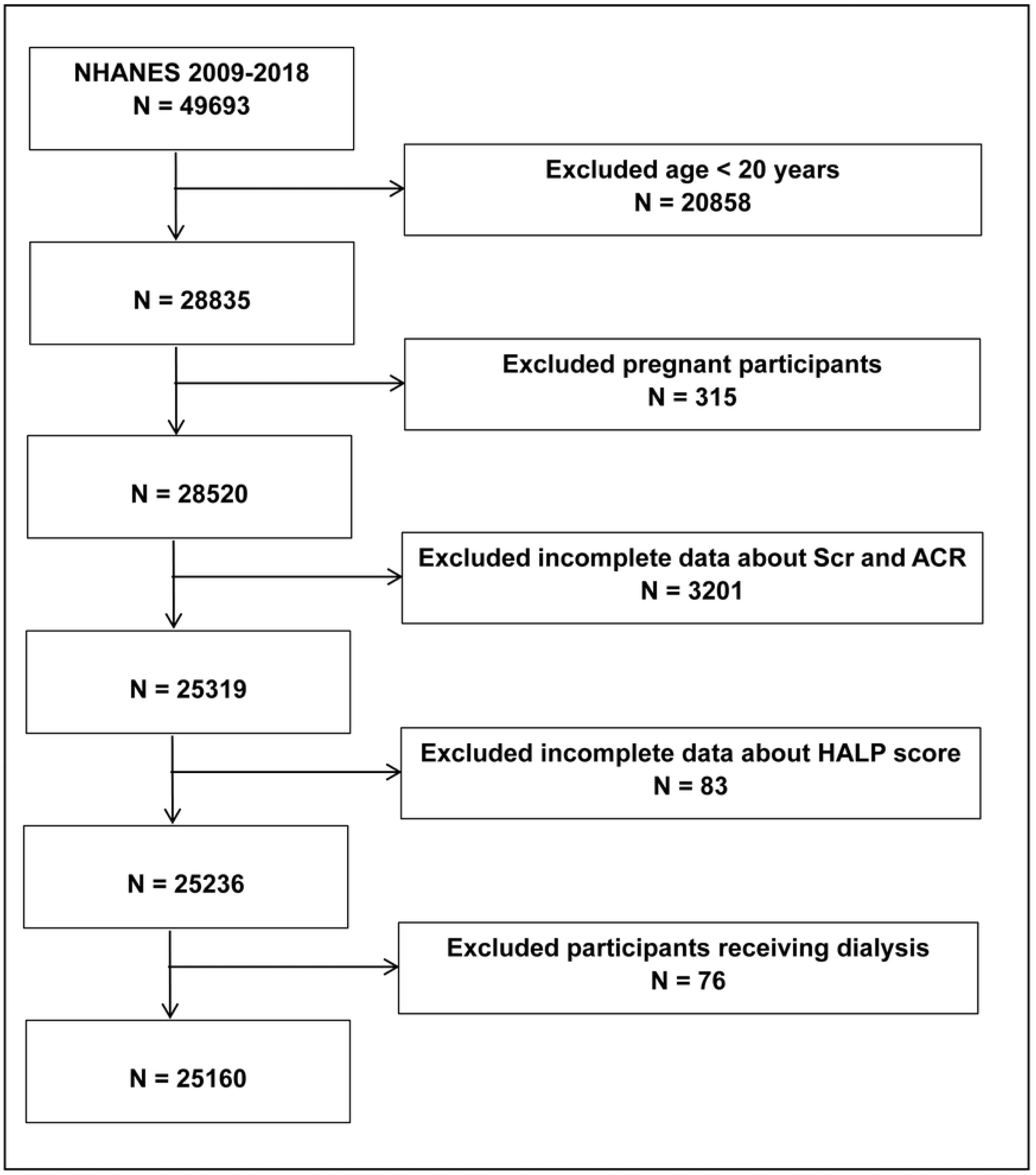
Flowchart of participants selection. Abbreviations: NHANES = National Health and Nutrition Examination Survey, Scr = serum creatinine, ACR = urine albumin-to-creatinine ratio, HALP score = hemoglobin, albumin, lymphocyte, and platelet score.

### 2.3 Assessment of HALP score

The HALP score was treated as the primary exposure variable and calculated as follows: HALP score = [hemoglobin (g/L) × serum albumin (g/L) × lymphocyte count (10⁹/L)] / platelet count (10⁹/L). Hematological parameters were determined through an automated blood cell analyzer (Beckman Coulter), while quantification of serum albumin relied on the bromocresol purple binding assay.

### 2.4 Assessment of CKD

CKD was determined based on albuminuria or a low-estimated glomerular filtration rate (eGFR). An eGFR < 60 mL/min/1.73 m² was considered low, with eGFR calculated using the Chronic Kidney Disease Epidemiology Collaboration (CKD-EPI) equation. Albuminuria corresponded to an ACR of ≥ 30 mg/g. CKD, albuminuria, and low-eGFR were considered as dependent variables in this study.

### 2.5 Covariates

Demographic variables included age (< 60 years or ≥ 60 years), gender (male or female), race (Mexican American, Other Hispanic, Non-Hispanic White, Non-Hispanic Black, and Other), poverty-to-income ratio (PIR; < 1, 1-4, or > 4), education levels (high school or less and more than high school), and marital status (married, widowed, divorced, separated, never married, or living with a partner). Physical and laboratory parameters considered were body mass index (BMI; < 25, 25-30, or ≥ 30 kg/m²), eGFR, ACR, blood urea nitrogen (BUN), Scr, serum uric acid (SUA), hemoglobin (HGB), lymphocytes (LYM), platelets (PLT), serum albumin (ALB), alanine transaminase (ALT), aspartate transaminase (AST), low-density lipoprotein cholesterol (LDL-C), high-density lipoprotein cholesterol (HDL-C), total cholesterol (TC), and triglycerides (TG).

Health-related behaviors and comorbidities were also incorporated. Smoking status was dichotomized as yes/no, defined by lifetime cigarette consumption ≥ 100 sticks. Drinking status was categorized as yes/no, based on annual alcoholic beverage intake exceeding 12 servings. Hypertension was defined as mean systolic blood pressure ≥ 140 mmHg, mean diastolic blood pressure ≥ 90 mmHg, self-reported hypertension, or current use of antihypertensive medications. Diabetes mellitus (DM) was identified by glycohemoglobin ≥ 6.5%, fasting glucose ≥ 7.0 mmol/L, a confirmed medical diagnosis of DM, or ongoing use of DM medications or insulin therapy. Participants who ever reported having congestive heart failure, coronary heart disease, angina pectoris or heart attack were diagnosed as having cardiovascular disease (CVD). Participants with TC ≥ 5.2 mmol/L, TG ≥ 1.7 mmol/L, LDL-C ≥ 3.4 mmol/L, HDL-C ≤ 1.0 mmol/L, self-reported hypercholesterolemia, or on lipid-lowering therapy were identified as having dyslipidemia.

### 2.6 Animal experimental design

#### 2.6.1 Experimental animals

All experimental procedures involving animals were approved by the Animal Ethics Committee of Hebei Provincial Hospital of Traditional Chinese Medicine (IACUC-HPHCM-2026015). Male Sprague-Dawley (SD) rats (200 ± 20 g body weight) were procured from Liaoning Changsheng Biotechnology Co., Ltd. Animals were maintained in specific pathogen-free (SPF) facilities at the Experimental Animal Center under controlled environmental conditions (22 ± 2 °C, 50 ± 10% humidity, 12-h photoperiod) with unrestricted access to standard diet and drinking water. Experiments were conducted in line with local animal welfare standards and the 3R principles to alleviate animal discomfort. All animal procedures were conducted by investigators certified in laboratory animal handling.

#### 2.6.2 CKD model establishment

After a one-week acclimatization period, rats were randomly allocated to the control (CON) group or the CKD model group (n = 10 per group). The CKD model was established using the two-step 5/6 nephrectomy procedure (Platt’s method) with minor modifications. [14] Briefly, rats were anesthetized via intraperitoneal injection of 1% pentobarbital sodium (50 mg/kg), with adequate anesthetic depth confirmed by the absence of the pedal withdrawal reflex prior to surgery. Following routine skin preparation and disinfection, a longitudinal incision was made at the left costovertebral angle to expose the left kidney. Approximately one-third of both the upper and lower poles of the renal cortex were resected separately, and hemostasis was achieved using gelatin sponges. The kidney was repositioned, and the incision was closed in layers. One week later, a total right nephrectomy was performed: the right renal pedicle was carefully ligated to prevent bleeding, and the kidney was excised intact before the abdominal cavity was closed in layers. Postoperative analgesia was provided by subcutaneous buprenorphine (0.05 mg/kg) at 12-h intervals for 48 h after each surgery, and animals were warmed on a heating pad until they had fully recovered from anesthesia.

Two weeks after the second surgery, successful CKD model establishment was confirmed by elevated serum Scr and BUN levels and characteristic renal histopathological changes. The total experimental duration was 4 weeks from the initial surgery (1 week between the two surgeries plus 2 weeks of post-modeling observation/treatment).

#### 2.6.3 Animal monitoring and humane endpoints

Throughout the entire study, the health and behavior of every animal were assessed no less than once per day, and body weight was documented on two occasions each week. Observation indicators comprised body weight, intake of food and water, body posture, level of activity, fur appearance, breathing pattern, and healing of the surgical incision. The pre-established criteria mandating immediate euthanasia were as follows: (1) a reduction in body weight of more than 20% relative to baseline; (2) failure to consume food or water; (3) marked respiratory difficulty; (4) a sustained moribund condition or lack of response to external stimuli; (5) manifestations of intense or refractory pain or distress (such as hunched posture, piloerection, or self-mutilation); or (6) wound rupture or severe infection that did not respond to veterinary treatment.

Of the 20 rats enrolled, all reached the scheduled study endpoint and were euthanized according to the planned protocol. No unexpected deaths occurred during the experiment, and none of the animals met the pre-established criteria that would have warranted earlier euthanasia.

#### 2.6.4 Euthanasia and sample collection

Following the 2-week post-modeling treatment phase, all rats were fasted overnight before being euthanized with isoflurane. In brief, anesthetic induction was achieved by exposing animals to 3–5% isoflurane in oxygen within an induction chamber, after which the concentration was lowered to 1.5–2.5% for maintenance until nociceptive reflexes were entirely abolished, as verified by the disappearance of both pedal withdrawal and corneal responses. Whole blood was subsequently obtained through abdominal aortic puncture under aseptic conditions, and serum was separated by centrifugation.

Throughout this procedure, isoflurane delivery was sustained until respiration and cardiac activity had completely ceased, thereby confirming death. Both kidneys were then aseptically excised and decapsulated, and a portion of each kidney was immersed in 4% paraformaldehyde for 48 h for subsequent histological evaluation.

#### 2.6.5 Biochemical analyses

Serum concentrations of BUN, Scr, and ALB were measured with commercial assay kits (Raydu Biotech, Wuhan, China) according to the manufacturer’s instructions, using the urease–glutamate dehydrogenase method (Cat. No. S036), an enzymatic assay (Cat. No. S03076), and the bromocresol green method (Cat. No. S03043), respectively. Urinary albumin and creatinine were determined in matched urine samples with the Rat Microalbuminuria ELISA Kit (Cat. No. E-EL-R0025) and the Creatinine Colorimetric Assay Kit (Cat. No. E-BC-K188-M; both from Elabscience, Wuhan, China), and the urinary albumin-to-creatinine ratio (UACR) was calculated accordingly. Hemoglobin (Hb), lymphocyte (LYM), and platelet (PLT) counts were determined on a Mindray BC-2800Vet veterinary hematology analyzer (Mindray, Shenzhen, China).

#### 2.6.6 Histopathological analysis

Continuous 4-μm sections underwent H&E and Masson trichrome staining for assessment of glomerular morphology, tubular injury, and interstitial fibrosis.

### 2.7 Statistical analysis

All analyses incorporated NHANES survey design and sampling weights to ensure national representativeness, in accordance with NHANES analytical guidelines. Data for continuous variables are presented as mean ± SD, and categorical data are summarized using proportions. To assess differences in baseline profiles across quartiles of the HALP score, weighted linear regression was applied to continuous measures, while weighted chi-square testing was employed for categorical ones. The relationships of the HALP score with CKD, albuminuria, and low-eGFR were investigated through weighted multivariable logistic regression, implemented across three sequential models. The crude model (Model 1) included no adjustments; Model 2 incorporated demographic factors including age, gender, and race; Model 3 additionally controlled for marital status, educational level, PIR, BMI, smoking and drinking, AST, ALT, hypertension, DM, CVD, and dyslipidemia. To explore potential non-linear associations, smooth curve fitting and generalized additive models (GAMs) were applied, followed by two-piecewise linear regression to identify threshold effects, with the inflection point determined by maximum likelihood estimation. Stratified analyses by age, gender, BMI, hypertension, and DM were performed, and interactions were assessed using likelihood ratio tests. For sensitivity analyses, Model 3 was rerun after excluding participants with HALP scores in the top and bottom 5% of the distribution. Comparative evaluation of diagnostic capability among the HALP score, PNI, PLR, LMR, and SII was performed via ROC curve analysis. To address missingness, median substitution was applied to continuous variables and mode substitution to categorical ones. Data processing relied on R software (version 4.1.3) and EmpowerStats, with significance defined at a two-sided threshold of P < 0.05.

For animal experiments, statistical analyses were performed using SPSS version 26.0. Data are expressed as mean ± standard error of the mean (SEM). Independent-samples t-tests were used to compare renal function indicators and HALP scores between the CON and CKD model groups. Pearson correlation analysis was performed to examine the associations between HALP score and serum BUN, Scr, and UACR concentrations in CKD rats.

## 3 Results

### 3.1 Baseline characteristics

A total of 25,160 individuals participated, with a mean age of 49.68 ± 17.54 years, comprising 48.94% males and 51.06% females. The overall prevalence of CKD was 17.30%. Age, gender, BMI, CKD, eGFR, ACR, BUN, Scr, SUA, HGB, LYM, PLT, ALB, ALT, AST, TG, and smoking status differed significantly across HALP score quartiles (all *P* < 0.05) (Table 1).

**Table 1.**
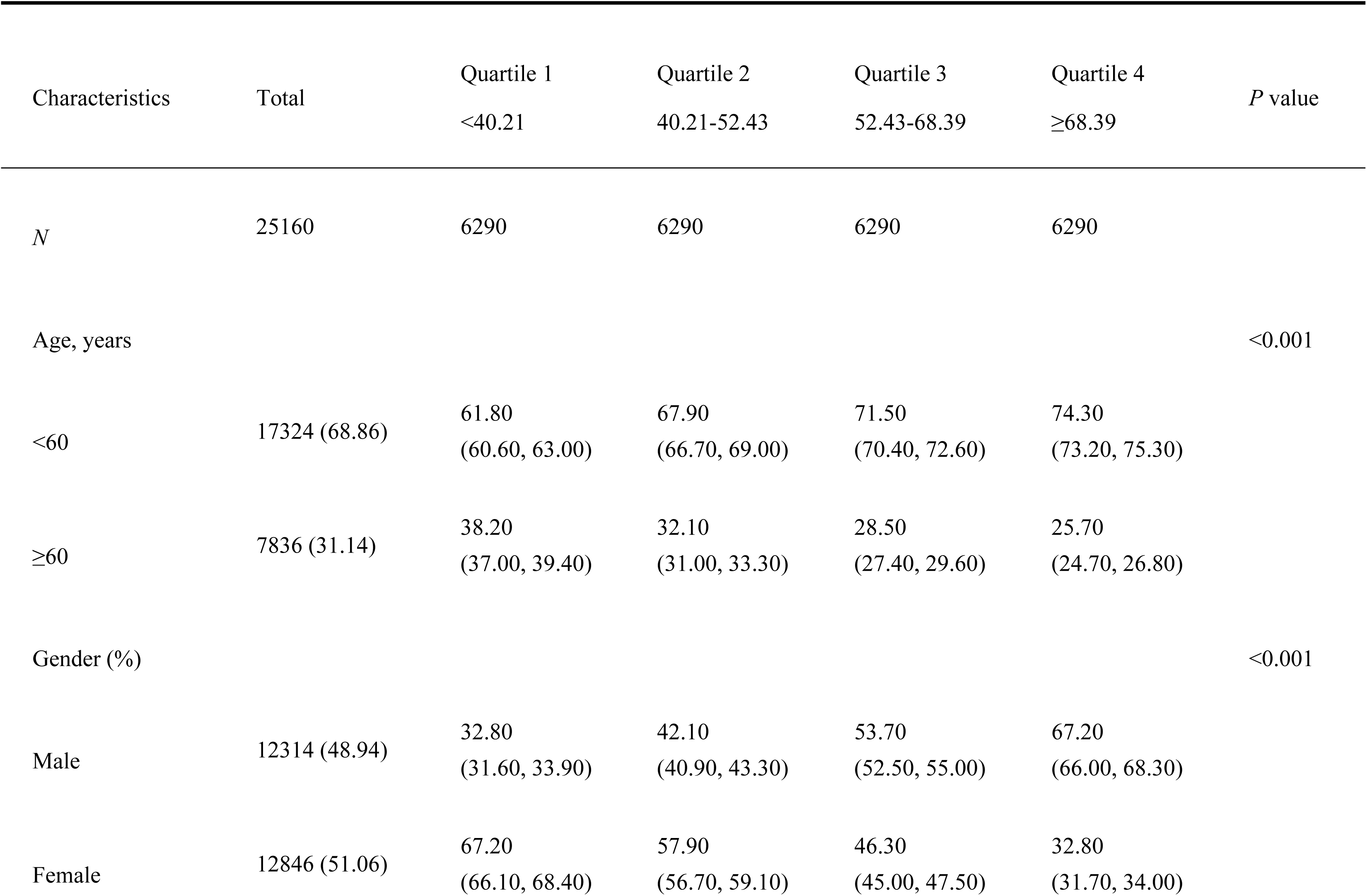
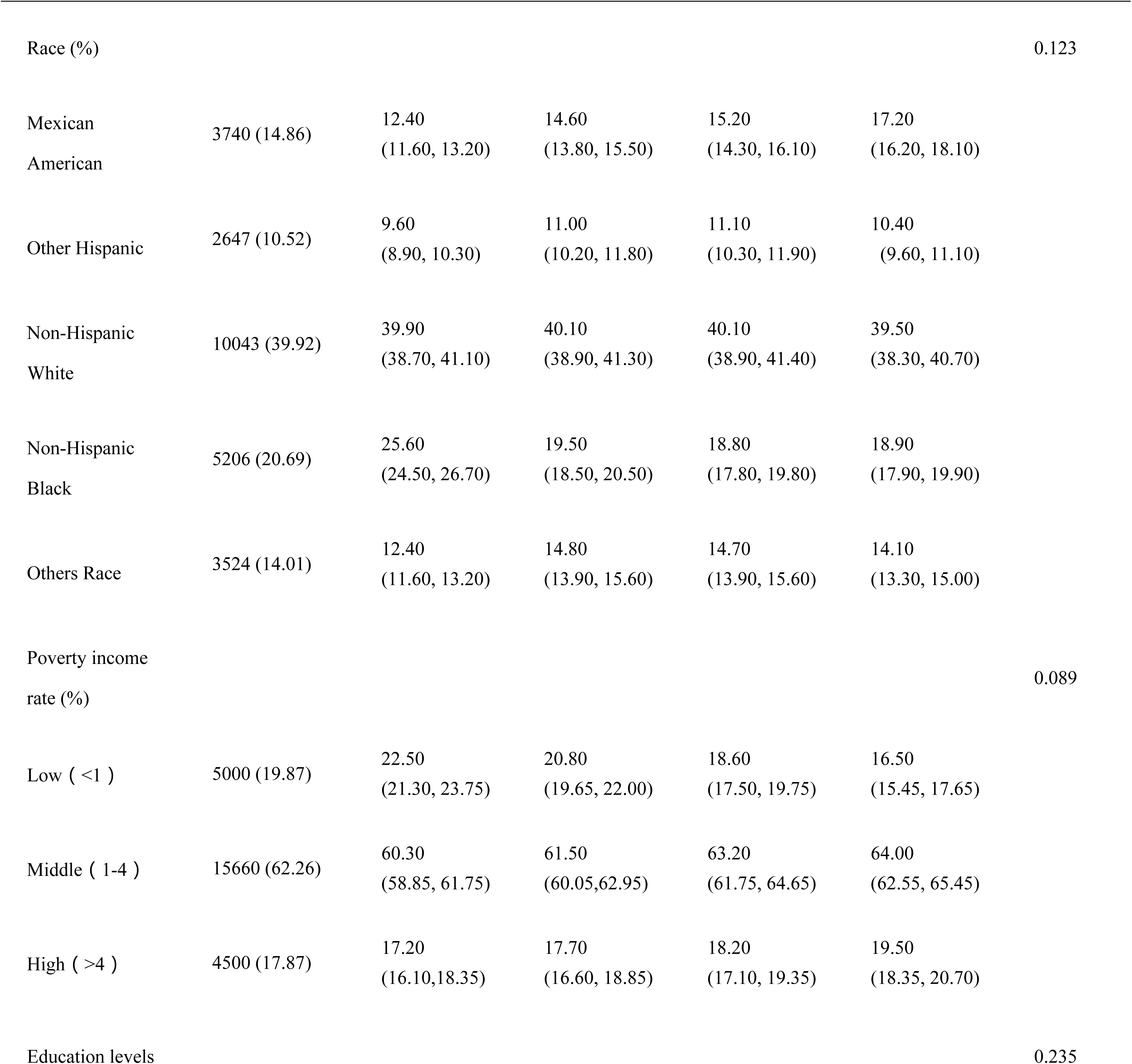
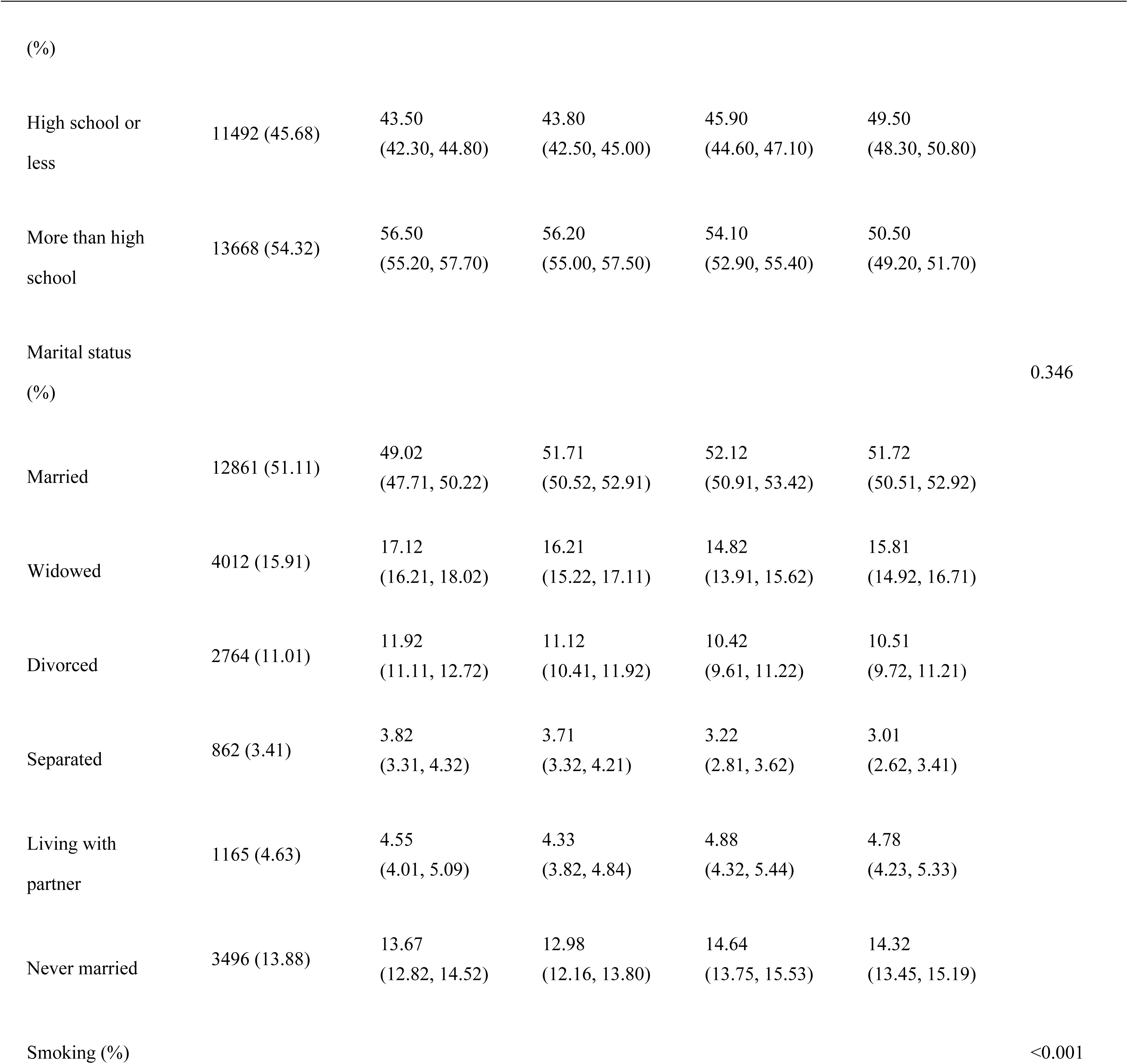
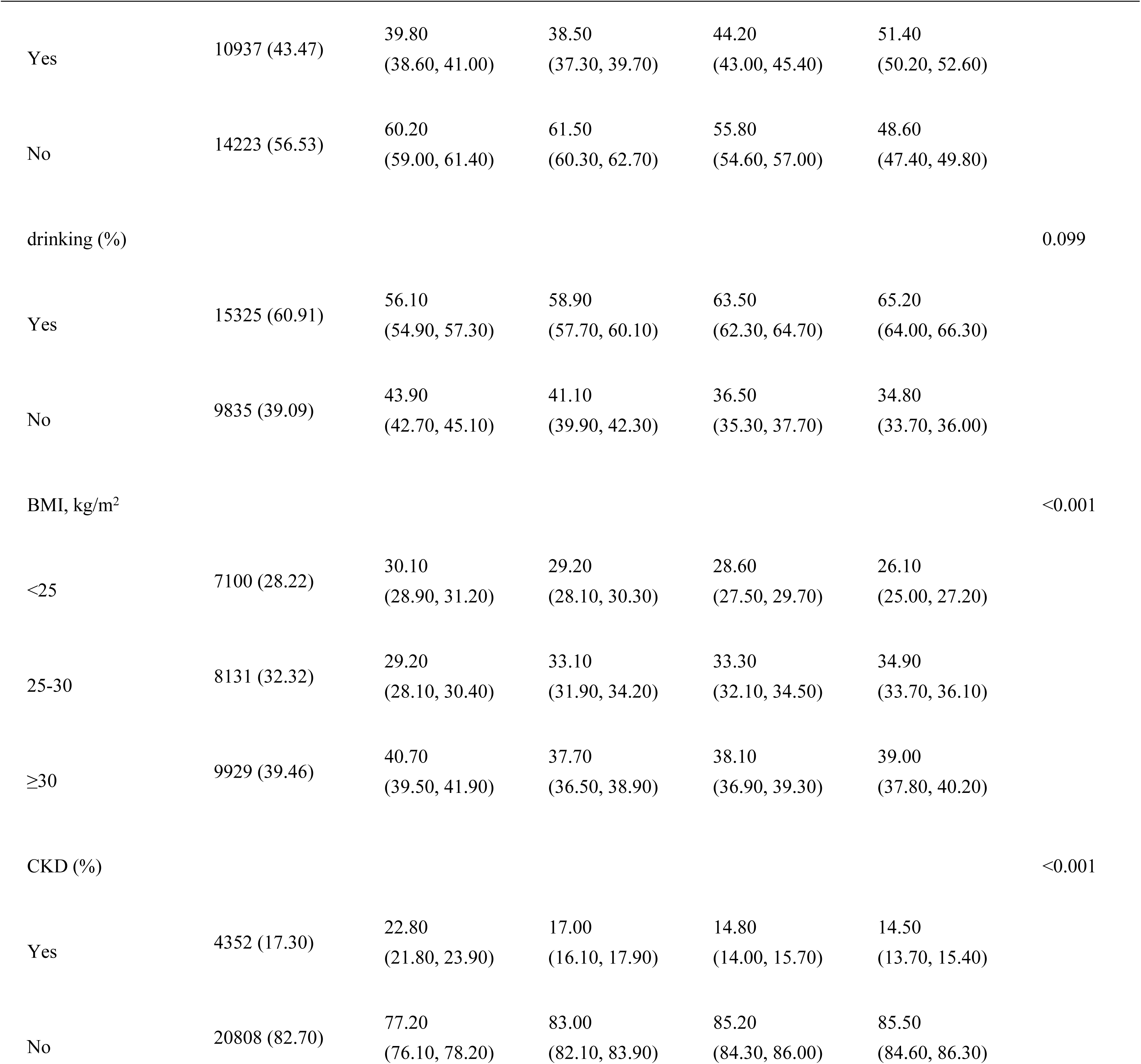
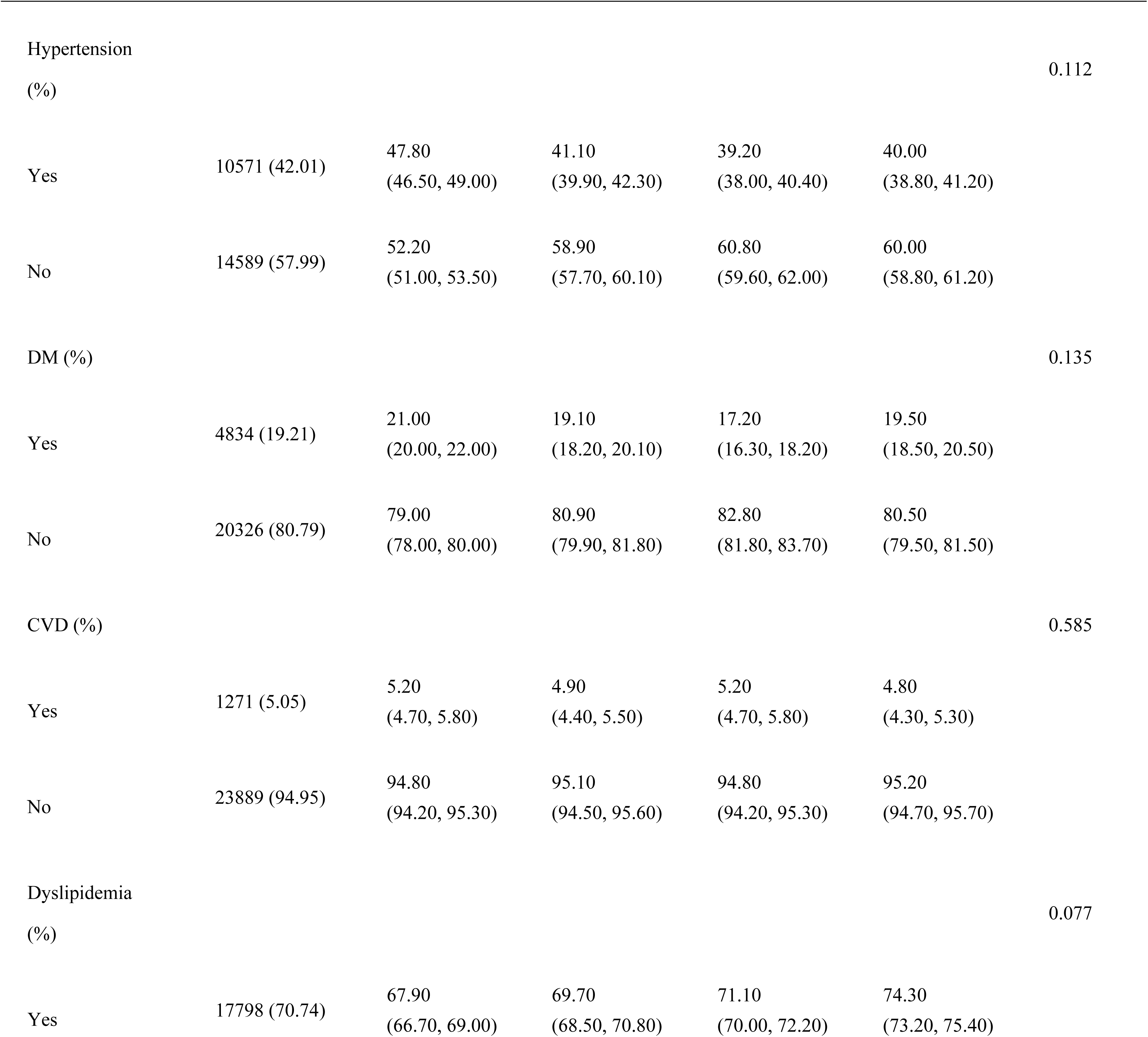
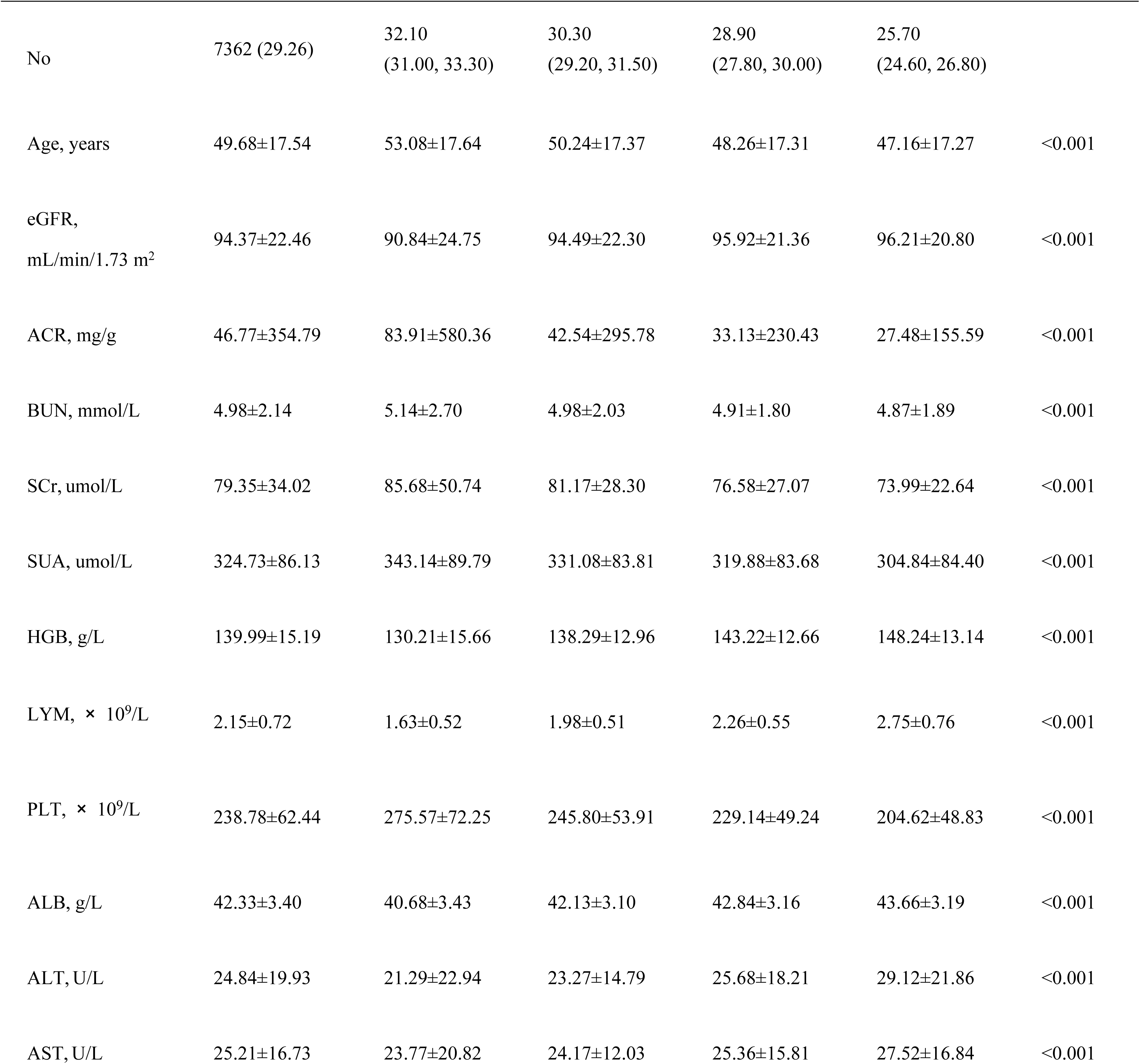
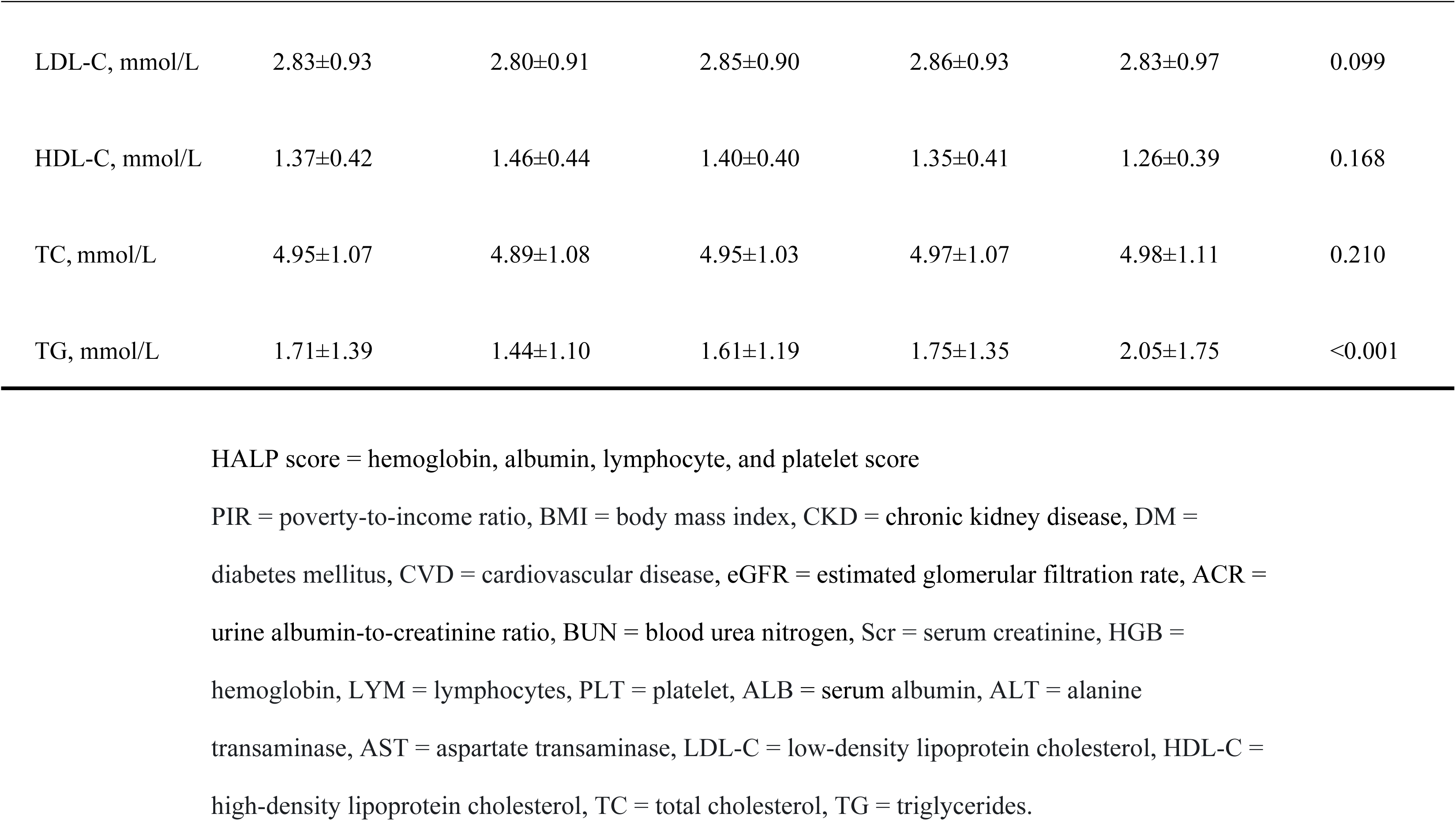
Baseline characteristics according to HALP score quartiles.

### 3.2 Relationship between HALP score and CKD, albuminuria, and low-eGFR

#### 3.2.1 Relationship between HALP score and CKD

As shown in Table 2, a higher HALP score was associated with reduced CKD prevalence in all models. Furthermore, when treated as a continuous variable, each additional point in the HALP score was associated with a 3% lower prevalence of CKD (OR = 0.97, 95% CI: 0.94–0.99, *P* = 0.031). Consistent results were observed in quartile-based analyses, with participants in the top quartile demonstrating a markedly reduced CKD risk compared with individuals in the bottom quartile in Model 3 (OR = 0.63, 95% CI: 0.52–0.76, *P* = 0.001).

**Table 2.** Associations between HALP score and CKD, albuminuria, and low-eGFR.

|  | Crude model (Model 1)<sup>3</sup> |  | Adjusted model (Model 2)<sup>4</sup> |  | Adjusted model (Model 3)<sup>5</sup> |  |
| --- | --- | --- | --- | --- | --- | --- |
|  | OR<sup>1</sup>(95%CI<sup>2</sup>) | *P* value | OR (95%CI) | *P* value | OR (95%CI) | *P* value |
| CKD |  |  |  |  |  |  |
| continuous | 0.95 (0.92-0.98) | 0.002 | 0.96 (0.93-0.99) | 0.016 | 0.97 (0.94-0.99) | 0.031 |
| Q1 | Reference |  | Reference |  | Reference |  |
| Q2 | 0.85 (0.75-0.96) | 0.010 | 0.86 (0.77-0.96) | 0.035 | 0.91(0.80-0.98) | 0.048 |
| Q3 | 0.70 (0.61-0.80) | <0.001 | 0.73 (0.63-0.84) | <0.001 | 0.78 (0.66-0.92) | 0.004 |
|  | OR<sup>1</sup>(95%CI<sup>2</sup>) | *P* value | OR (95%CI) | *P* value | OR (95%CI) | *P* value |
| Q4 | 0.55 (0.47-0.64) | <0.001 | 0.58 (0.49-0.68) | <0.001 | 0.63 (0.52-0.76) | 0.001 |
| *P* for trend | <0.001 |  | <0.001 |  | 0.008 |  |
| Albuminuria |  |  |  |  |  |  |
| continuous | 0.94 (0.91-0.97) | <0.001 | 0.95 (0.92-0.98) | 0.003 | 0.97 (0.93-0.99) | 0.044 |
| Q1 | Reference |  | Reference |  | Reference |  |
| Q2 | 0.82 (0.72-0.93) | 0.003 | 0.85 (0.74-0.97) | 0.017 | 0.89 (0.79-1.09) | 0.095 |
| Q3 | 0.68 (0.59-0.78) | < 0.001 | 0.71 (0.61-0.82) | < 0.001 | 0.75 (0.64-0.88) | 0.001 |
| Q4 | 0.52 (0.44-0.61) | < 0.001 | 0.55 (0.46-0.65) | < 0.001 | 0.60 (0.49-0.73) | 0.001 |
| *P* for trend | < 0.001 |  | <0.001 |  | 0.006 |  |
| Low-eGFR |  |  |  |  |  |  |
| continuous | 0.96 (0.93-0.99) | 0.015 | 0.97 (0.94-0.99) | 0.021 | 0.98 (0.95-1.01) | 0.065 |
| Q1 | Reference |  | Reference |  | Reference |  |
| Q2 | 0.89 (0.81-0.98) | 0.020 | 0.91 (0.82-0.99) | 0.042 | 0.95 (0.85-1.06) | 0.089 |
| Q3 | 0.85 (0.76-0.95) | 0.007 | 0.88 (0.79-0.98) | 0.022 | 0.91 (0.81-1.02) | 0.078 |
| Q4 | 0.78 (0.70-0.87) | 0.002 | 0.81 (0.73-0.90) | 0.008 | 0.84 (0.75-1.01) | 0.055 |
| *P* for trend | 0.005 |  | 0.018 |  | 0.062 |  |

<sup>1</sup>OR: Odd ratio
<sup>2</sup> 95% CI: 95% confidence interval
<sup>3</sup> Model 1: No covariates were adjusted
<sup>4</sup> Model 2: Adjusted for age, gender, and race
<sup>5</sup> Model 3: Adjusted for age, gender, race, marital status, education level, PIR, BMI, smoking, drinking, AST, ALT, hypertension, DM, CVD, and dyslipidemia.
HALP score = hemoglobin, albumin, lymphocyte, and platelet score, CKD = chronic kidney disease, eGFR = estimated glomerular filtration rate, PIR = poverty-to-income ratio, BMI = body mass index, AST = aspartate transaminase, ALT = alanine transaminase, DM = diabetes mellitus, CVD = cardiovascular disease.

Smooth curve fitting further indicated a non-linear connection between the HALP score and CKD risk (Fig 2A). Specifically, CKD prevalence decreased as the HALP score increased, reaching its lowest point at a HALP score threshold of 52.43 (OR = 0.97, 95% CI: 0.95–0.99, *P* < 0.05). Beyond this threshold, further increases in the HALP score conferred no additional statistically significant protection (OR = 1.00, 95% CI: 0.98–1.01, *P* = 0.786) (Table 3).

**Fig 2.**
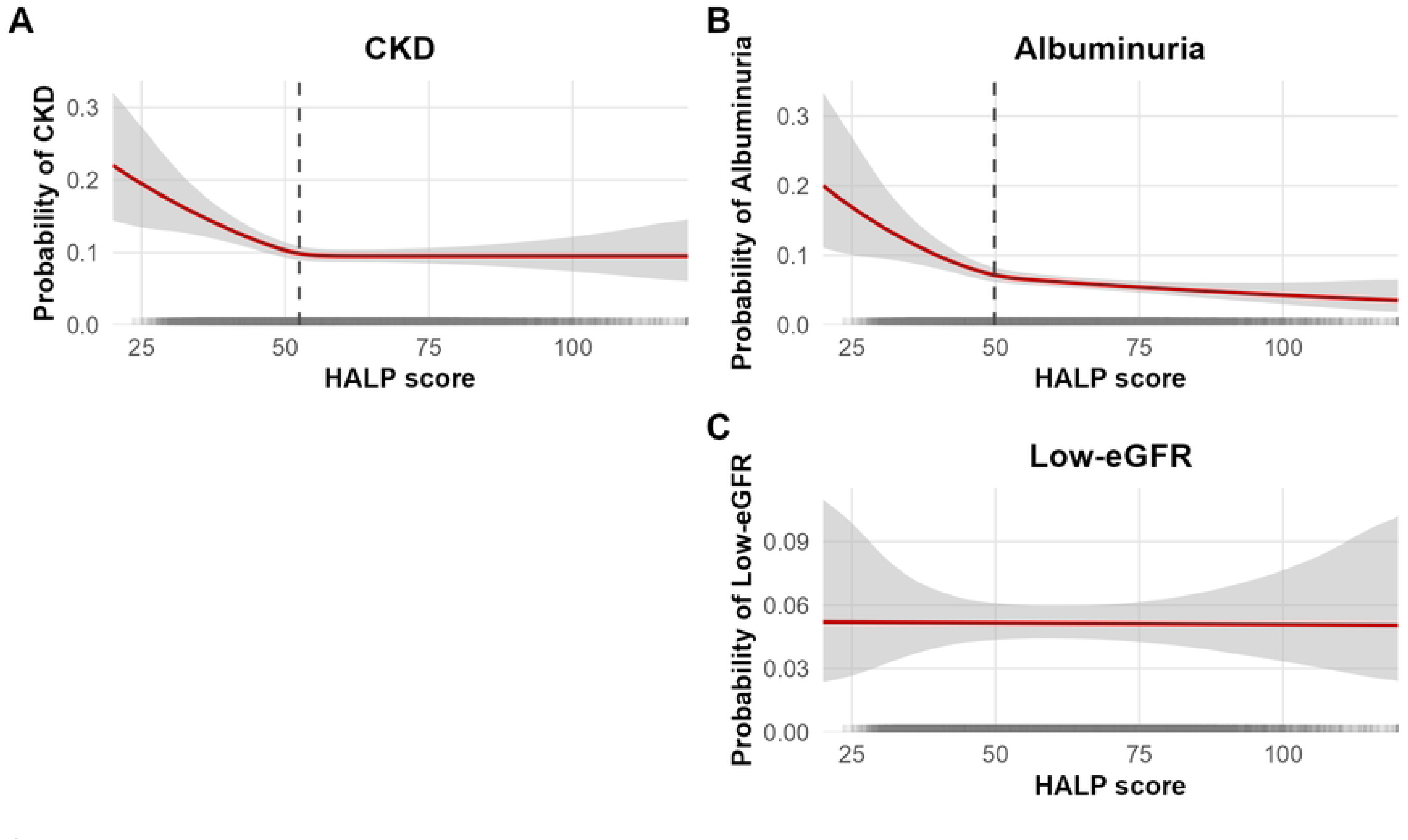
Smooth curve fitting depicting the non-linear associations of the HALP score with CKD (A), albuminuria (B), and low-eGFR (C), assessed using generalized additive models. The solid lines represent the fitted curves, and the shaded areas represent the 95% confidence intervals. A HALP score and CKD; B HALP score and albuminuria; C HALP score and low-eGFR Abbreviations: CKD = chronic kidney disease, eGFR = estimated glomerular filtration rate, HALP score = hemoglobin, albumin, lymphocyte, and platelet score.

**Table 3.** Threshold effect analysis of the HALP score on CKD using a two-piecewise linear regression model after adjustment of covariates.

| CKD | Adjusted OR<sup>1</sup> (95%CI<sup>2</sup>) | *P* value |
| --- | --- | --- |
| Fitting by the standard linear model | 0.97 (0.94-0.99) | 0.032 |
| Fitting by the two-piecewise linear model |  |  |
| HALP score |  |  |
| Inflection point | 52.43 |  |
| HALP score < 52.43 | 0.97 (0.95-0.99) | 0.023 |
| HALP score ≥ 52.43 | 1.00 (0.98-1.01) | 0.786 |
| Log likelihood ratio |  | 0.031 |

<sup>1</sup>OR: Odd ratio
<sup>2</sup> 95% CI: 95% confidence interval
Adjusted for age, gender, race, marital status, education level, PIR, BMI, smoking, drinking, AST, ALT, hypertension, DM, CVD, and dyslipidemia.
HALP score = hemoglobin, albumin, lymphocyte, and platelet score, CKD = chronic kidney disease, PIR = poverty-to-income ratio, BMI = body mass index, AST = aspartate transaminase, ALT = alanine transaminase, DM = diabetes mellitus, CVD = cardiovascular disease.

#### 3.2.2 Relationship between HALP score and albuminuria

A lower HALP score was similarly associated with a higher prevalence of albuminuria (OR = 0.97, 95% CI: 0.93–0.99, *P* = 0.044). This association persisted when the HALP score was categorized into quartiles, with participants in lower quartiles showing a higher prevalence of albuminuria compared with those in higher quartiles (trend test, *P* < 0.05) (Table 2).

Smooth curve fitting revealed a non-linear, logarithmic-like relationship between the HALP score and albuminuria, with a consistent threshold across models (Fig 2B). Prior to the threshold, every single-point rise in HALP score was associated with a 4% lower prevalence of albuminuria (OR = 0.96, 95% CI: 0.94–0.98, *P* < 0.05), whereas above the threshold, further increases conferred no significant effect (OR = 0.99, 95% CI: 0.97–1.01, *P* = 0.352) (Table 4).

**Table 4.**
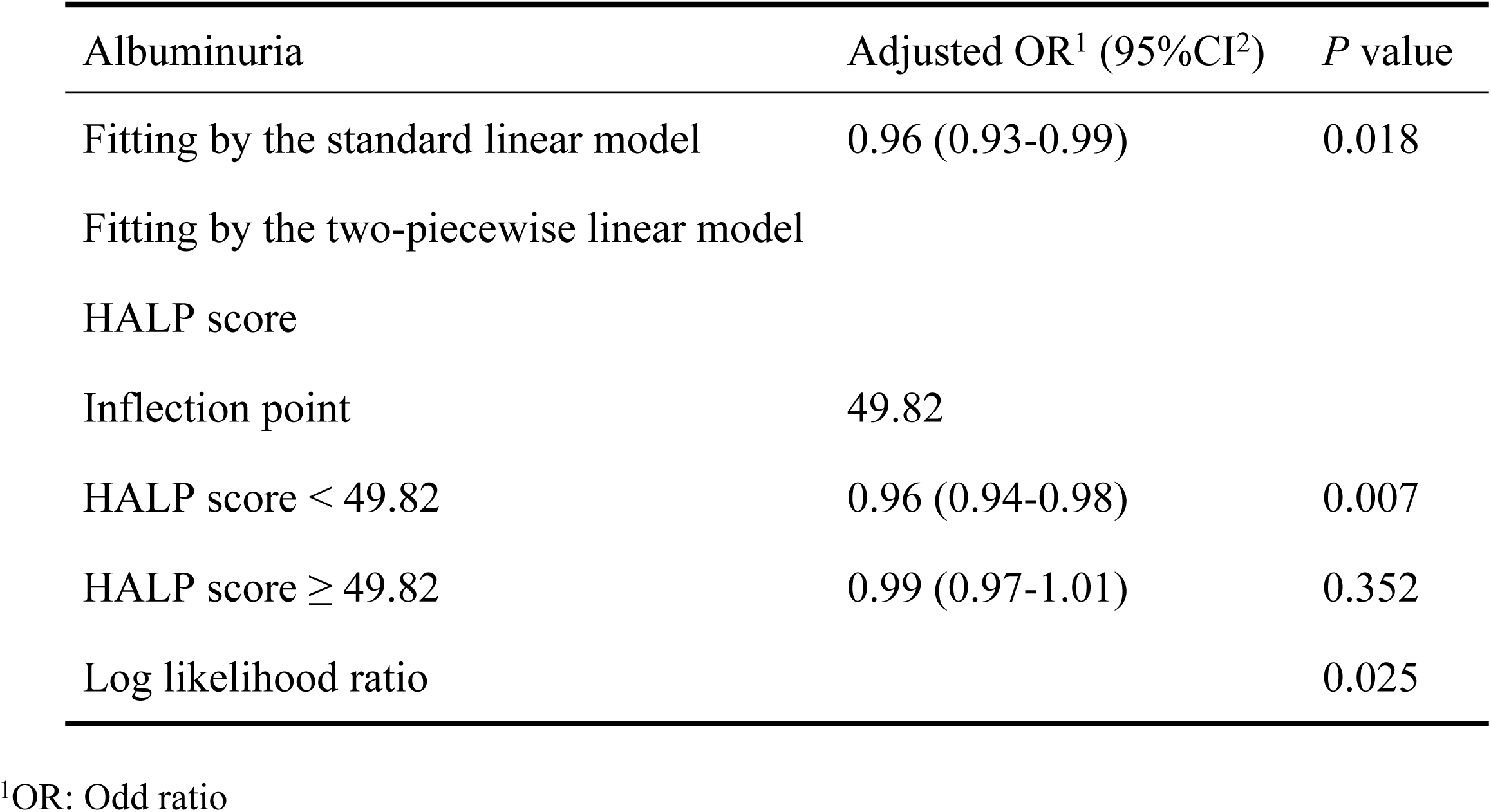
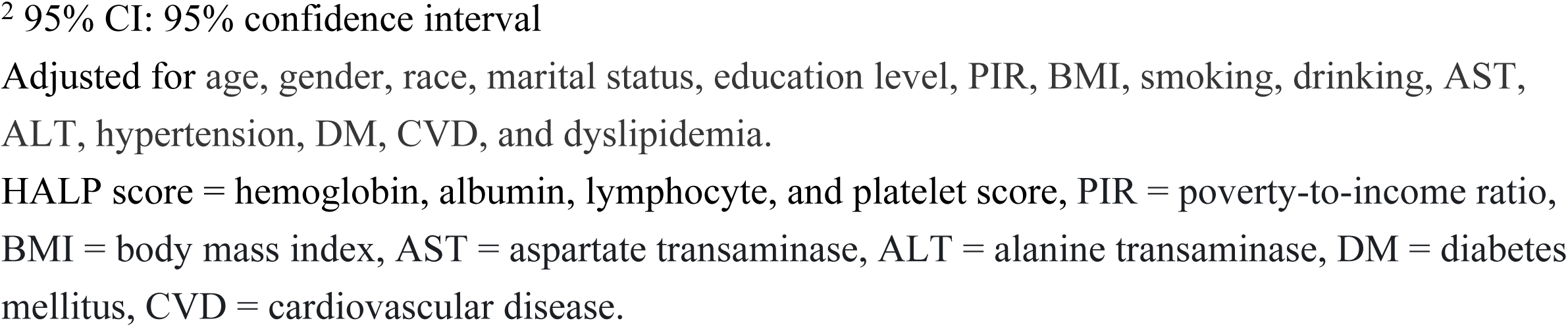
Threshold effect analysis of the HALP score on albuminuria using a two-piecewise linear regression model after adjustment of covariates.

| Albuminuria | Adjusted OR<sup>1</sup> (95%CI<sup>2</sup>) | *P* value |
| --- | --- | --- |
| Fitting by the standard linear model | 0.96 (0.93-0.99) | 0.018 |
| Fitting by the two-piecewise linear model |  |  |
| HALP score |  |  |
| Inflection point | 49.82 |  |
| HALP score < 49.82 | 0.96 (0.94-0.98) | 0.007 |
| HALP score ≥ 49.82 | 0.99 (0.97-1.01) | 0.352 |
| Log likelihood ratio |  | 0.025 |

<sup>1</sup>OR: Odd ratio
<sup>2</sup> 95% CI: 95% confidence interval
Adjusted for age, gender, race, marital status, education level, PIR, BMI, smoking, drinking, AST, ALT, hypertension, DM, CVD, and dyslipidemia.
HALP score = hemoglobin, albumin, lymphocyte, and platelet score, PIR = poverty-to-income ratio, BMI = body mass index, AST = aspartate transaminase, ALT = alanine transaminase, DM = diabetes mellitus, CVD = cardiovascular disease.

#### 3.2.3 Relationship between HALP score and low-eGFR

In Model 1, the HALP score showed a significant inverse association with low-eGFR (OR = 0.96, 95% CI: 0.93–0.99, *P* < 0.05), and this association persisted in Model 2 (OR = 0.97, 95% CI: 0.94–0.99, *P* < 0.05). However, in Model 3, the association lost statistical significance (OR = 0.98, 95% CI: 0.95–1.01, *P* = 0.065) (Table 2). Furthermore, smooth curve fitting did not reveal a significant non-linear association between the HALP score and low-eGFR (Fig 2C).

### 3.3 Subgroup analysis

Subgroup analyses stratified by age, gender, hypertension, DM, and BMI were performed. Interaction analyses indicated that none of the stratification variables significantly modified the association between the HALP score and CKD (all *P* > 0.05). Similarly, the associations of the HALP score with albuminuria and low-eGFR remained stable across all evaluated subgroups (Fig 3).

**Fig 3.**
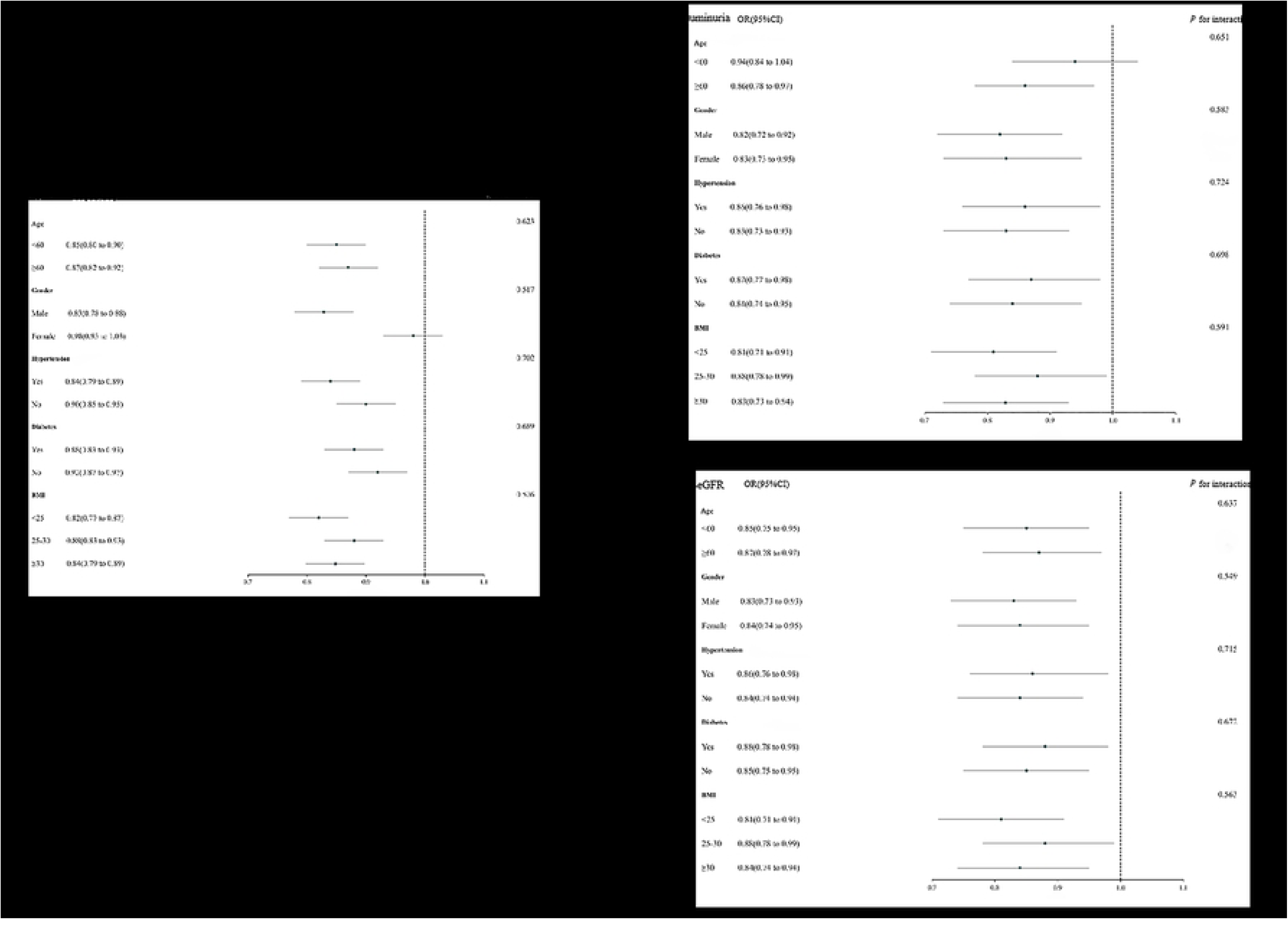
Subgroup analysis of the associations between the HALP score and CKD, albuminuria, and low-eGFR. A HALP score and CKD; B HALP score and albuminuria; C HALP score and low-eGFR Abbreviations: CKD = chronic kidney disease, eGFR = estimated glomerular filtration rate, HALP score = hemoglobin, albumin, lymphocyte, and platelet score.

### 3.4 Sensitivity analysis

To verify robustness of our findings, we performed sensitivity analyses by excluding participants with extreme HALP scores (lowest and highest 5%). The fully adjusted Model 3 was reapplied, and the results were consistent with the main analyses, confirming the stability of the inverse association between the HALP score and CKD (OR = 0.97, 95% CI: 0.94–0.99, *P* < 0.05) (Table 5).

**Table 5.**
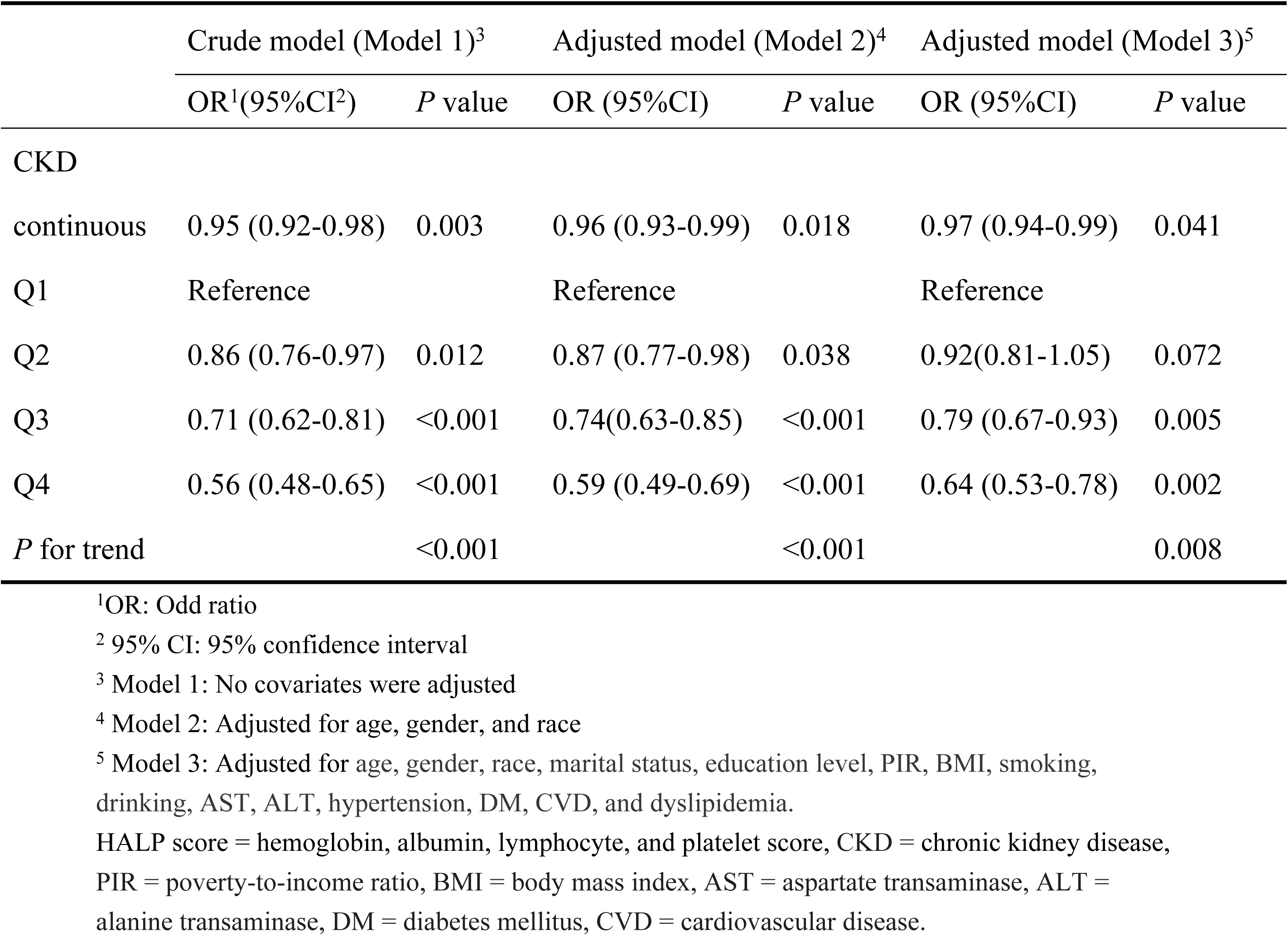
Sensitivity analysis after excluding extreme 5% of HALP score.

### 3.5 ROC analysis

ROC curve analyses were performed to compare the discriminative ability of the HALP score with other biomarkers (PNI, PLR, LMR, and SII) for CKD, albuminuria, and low-eGFR (Fig 4). The results revealed that HALP scores had significantly higher AUC values than other biomarkers, with significant differences for CKD and albuminuria (all *P* < 0.05, Table 6). These findings suggest that the HALP score offers superior discriminative ability for identifying CKD and albuminuria compared with other inflammation- and nutrition-related biomarkers.

**Fig 4.**
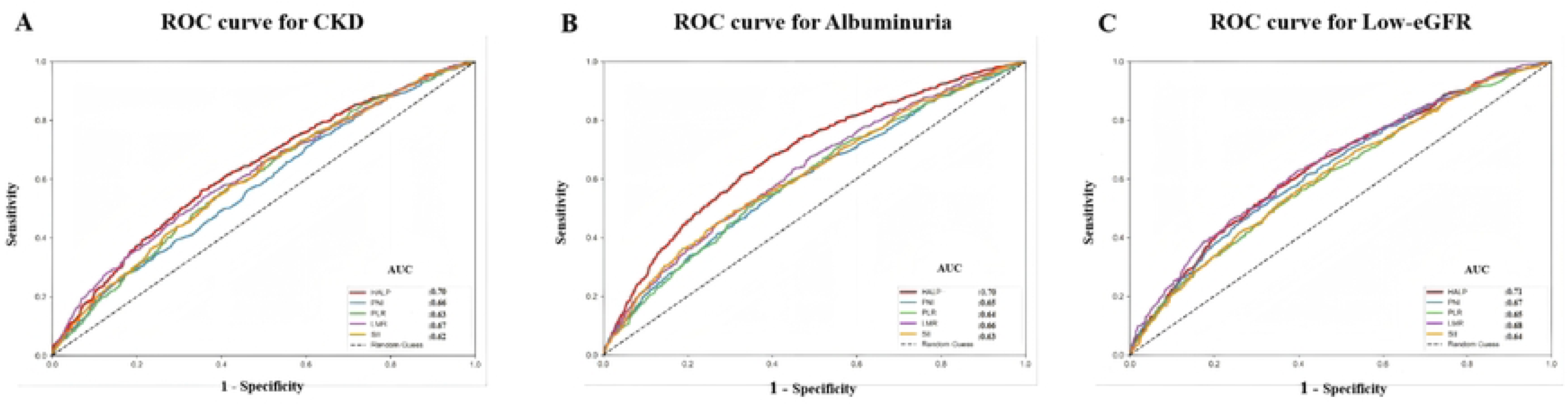
ROC curves and AUC values of the five diagnostic markers (HALP score, PNI, PLR, LMR, and SII) for CKD, albuminuria and low-eGFR. Five markers were assessed to identify CKD; B Five markers were assessed to identify albuminuria; C Five markers were assessed to identify low-eGFR Abbreviations: ROC = receiver operating characteristic, AUC = area under the curve, CKD = chronic kidney disease, eGFR = estimated glomerular filtration rate, HALP score = hemoglobin, albumin, lymphocyte, and platelet score, PNI = prognostic nutritional index, LMR = lymphocyte-to-monocyte ratio, PLR = platelet-to-lymphocyte ratio, SII = systemic immune-inflammation index.

**Table 6.** Comparison of AUC values between HALP score and other markers for predicting CKD, albuminuria, and low-eGFR.

| Test | AUC<sup>1</sup> | 95% CI<sup>2</sup>low | 95% CI upp | Best threshold | Specificity | Sensitivity | P for different in AUC |
| --- | --- | --- | --- | --- | --- | --- | --- |
| CKD |  |  |  |  |  |  |  |
| HALP score | 0.70 | 0.68 | 0.72 | 52.34 | 0.65 | 0.70 | Reference |
| PNI | 0.66 | 0.64 | 0.68 | 42.15 | 0.58 | 0.62 | <0.001 |
| PLR | 0.63 | 0.61 | 0.65 | 155.78 | 0.55 | 0.58 | <0.001 |

| Test | AUC<sup>1</sup> | 95% CI<sup>2</sup>low | 95% CI upp | Best threshold | Specificity | Sensitivity | *P* for different in AUC |
| --- | --- | --- | --- | --- | --- | --- | --- |
| LMR | 0.67 | 0.65 | 0.69 | 3.89 | 0.62 | 0.66 | <0.001 |
| SII | 0.62 | 0.60 | 0.64 | 610.23 | 0.52 | 0.56 | <0.001 |
| Albuminuria |  |  |  |  |  |  |  |
| HALP score | 0.70 | 0.67 | 0.71 | 51.23 | 0.50 | 0.75 | Reference |
| PNI | 0.65 | 0.63 | 0.67 | 41.34 | 0.46 | 0.72 | <0.001 |
| PLR | 0.64 | 0.62 | 0.66 | 154.89 | 0.58 | 0.56 | <0.001 |
| LMR | 0.66 | 0.64 | 0.68 | 3.78 | 0.42 | 0.70 | <0.001 |
| SII | 0.63 | 0.61 | 0.65 | 605.34 | 0.38 | 0.68 | <0.001 |
| Low-eGFR |  |  |  |  |  |  |  |
| HALP score | 0.71 | 0.69 | 0.73 | 53.45 | 0.66 | 0.52 | Reference |
| PNI | 0.67 | 0.65 | 0.69 | 43.21 | 0.52 | 0.63 | <0.001 |
| PLR | 0.65 | 0.63 | 0.67 | 156.78 | 0.59 | 0.55 | <0.001 |
| LMR | 0.68 | 0.66 | 0.70 | 3.90 | 0.61 | 0.54 | 0.85 |
| SII | 0.64 | 0.62 | 0.66 | 615.45 | 0.59 | 0.55 | <0.001 |

<sup>1</sup>AUC: area under the curve
<sup>2</sup>95% CI: 95% confidence interval
HALP score = hemoglobin, albumin, lymphocyte, and platelet score, CKD = chronic kidney disease, eGFR = estimated glomerular filtration rate, PNI = prognostic nutritional index, PLR = platelet-to-lymphocyte ratio LMR = lymphocyte-to-monocyte ratio, SII = systemic immune-inflammation index.

### 3.6 Experimental validation using a subtotal nephrectomy rat model

A reliable CKD rat model was successfully established. In the CON group, H&E staining showed normal glomerular morphology with regularly arranged capillary loops, and no obvious mesangial proliferation or matrix accumulation was observed. Renal tubules were regularly organized with intact epithelial cells and clear lumens, and no inflammatory infiltration or interstitial fibrosis was observed. By contrast, in the model group, glomeruli exhibited mesangial cell proliferation and increased matrix deposition, accompanied by disordered capillary loops, thickened Bowman’s capsule, and collapsed capillary tufts. Renal tubules were disordered with irregular luminal dilation, along with varying degrees of tubular epithelial degeneration, necrosis, and shedding. Marked inflammatory cell infiltration was also detected in the renal interstitium. Masson trichrome staining demonstrated widespread deposition of blue-stained collagen fibers in the model group, including glomerular mesangial areas, peri-Bowman’s capsule regions, and the renal interstitium, indicating interstitial fibrosis (Fig 5).

**Fig 5.**
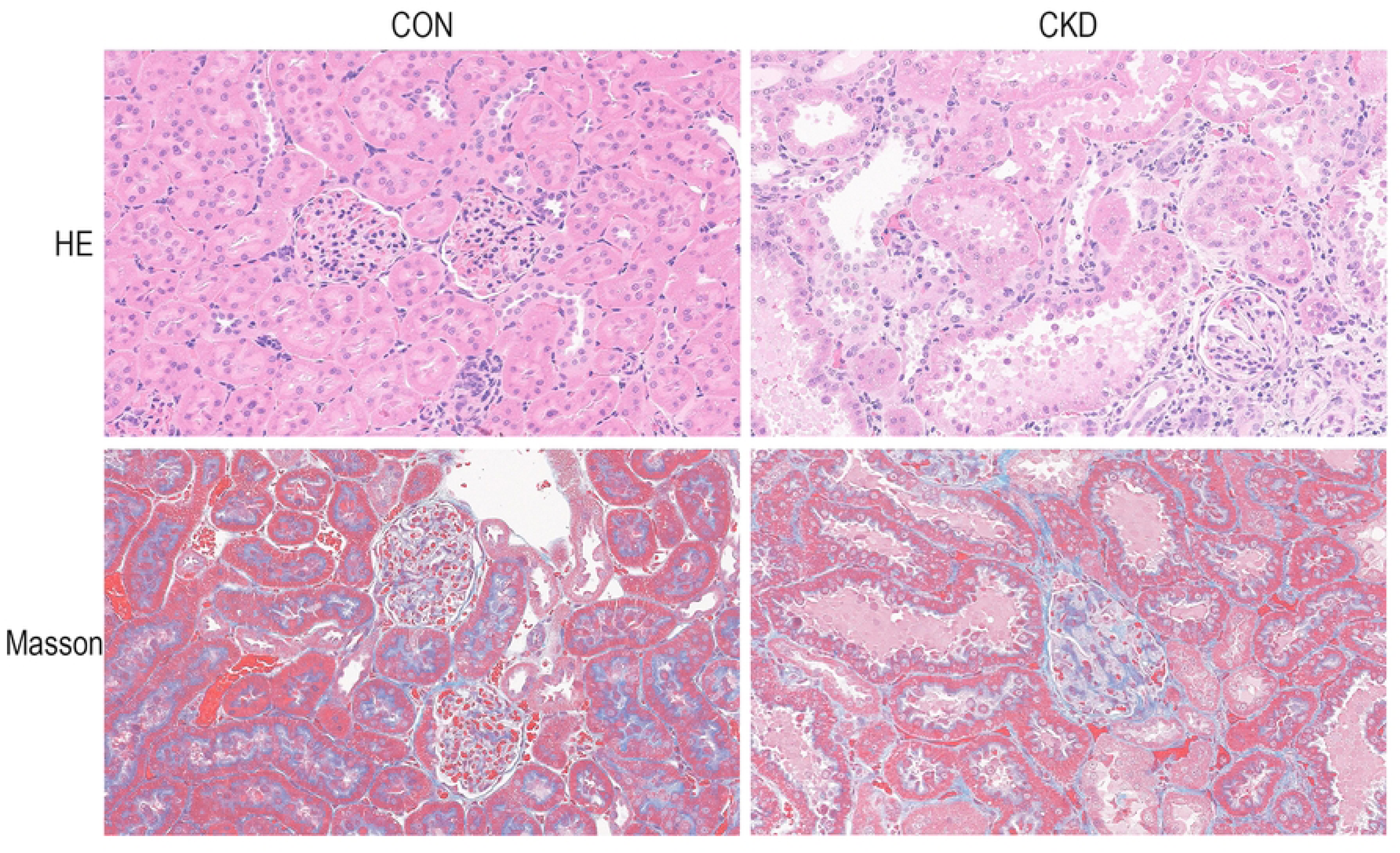
Representative HE and Masson staining of rat renal tissues. Scale bars: 50 μm.

Compared with the CON group, CKD model rats exhibited significantly elevated serum BUN, Scr, and UACR levels (all *P* < 0.05, Table 7, Fig 6). Meanwhile, CKD rats presented a markedly decreased HALP score, accompanied by reduced Hb, ALB, and LYM levels and increased PLT counts (all *P* < 0.05, Table 7, Fig 6).

**Fig 6.**
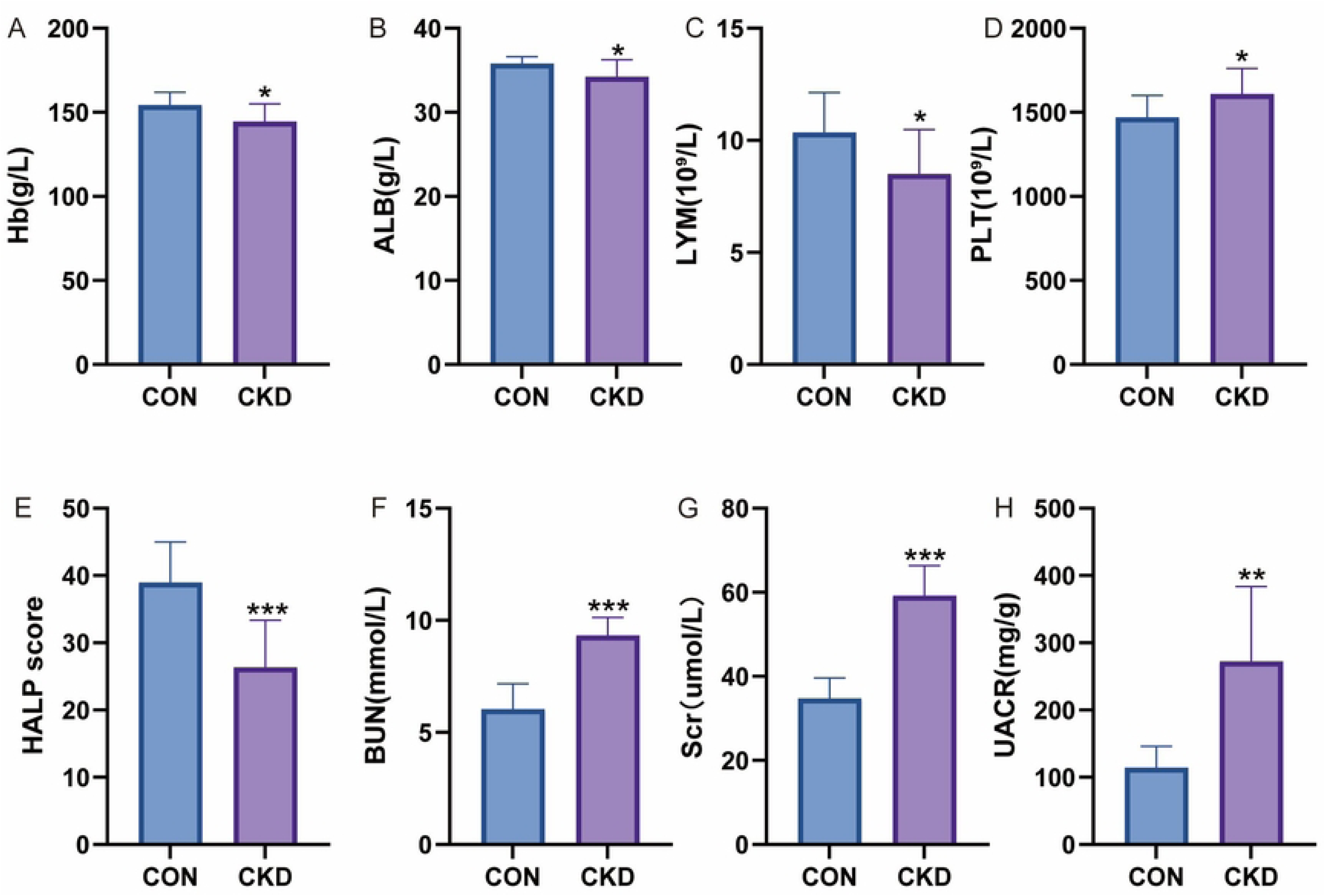
Comparison of hematological and renal function-related parameters between the control CON and CKD groups (A)Hb; (B) ALB; (C) LYM; (D) PLT; (E) HALP score; (F) BUN; (G) Scr; (H) UACR. Data are presented as mean ± SEM; *P<0.05, **P<0.01, ***P<0.001 vs. the CON group. HB = Hemoglobin, ALB = Albumin, LYM = lymphocytes, PLT = Platelet, HALP score = Hemoglobin, Albumin, Lymphocyte and Platelet score, BUN = Blood Urea Nitrogen, Scr = serum creatinine, UACR = Urinary Albumin-to-Creatinine Ratio

**Table 7.**
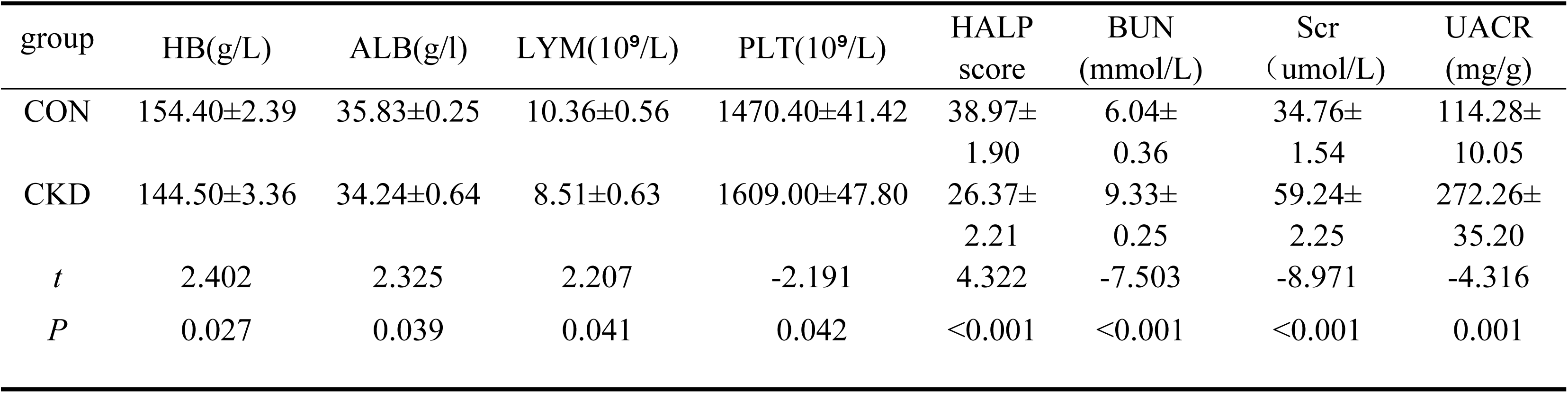
Comparison of HB, ALB, LYM, PLT, HALP score, BUN, Scr, and UACR levels in rat models (x±s,n= 10) HB = Hemoglobin, ALB = Albumin, LYM = lymphocytes, PLT = Platelet, HALP score = Hemoglobin, Albumin, Lymphocyte and Platelet score, BUN = Blood Urea Nitrogen, Scr = serum creatinine, UACR = Urinary Albumin-to-Creatinine Ratio

### 3.7 Correlation between HALP score and renal function indicators

Pearson correlation analysis demonstrated that the HALP score was significantly negatively correlated with serum BUN, Scr, and UACR levels in rats. Notably, UACR showed the strongest correlation with the HALP score (r = -0.771, *P* < 0.001, Table 8).

**Table 8.** Correlation of HALP score with renal function parameters in rats.

| Parameter | *r* | *P* |
| --- | --- | --- |
| BUN | -0.636 | 0.003 |
| Scr | -0.639 | 0.002 |
| UACR | -0.771 | <0.001 |

HALP score = Hemoglobin, Albumin, Lymphocyte and Platelet score, BUN = Blood Urea Nitrogen, Scr = serum creatinine, UACR = Urinary Albumin-to-Creatinine Ratio

## 4 Discussion

In this study, we found that a higher HALP score was independently and inversely associated with the prevalence of CKD and albuminuria in a representative U.S. adult cohort, whereas its association with low-eGFR was attenuated after full adjustment. A non-linear, threshold-type relationship was identified, with CKD risk reaching its nadir at a HALP score of 52.43; a comparable threshold pattern was also observed for albuminuria. These associations remained robust across subgroups stratified by age, sex, BMI, hypertension, and diabetes, and were preserved in sensitivity analyses. ROC analysis further demonstrated that the HALP score outperformed PNI, SII, LMR, and PLR in identifying CKD and albuminuria. Importantly, the clinical observations were corroborated in a 5/6 nephrectomy rat model, in which CKD rats exhibited markedly lower HALP scores that correlated negatively with serum BUN, Scr, and UACR, with UACR showing the strongest correlation.

CKD represents a significant worldwide health challenge, with its prevalence steadily increasing and imposing substantial burdens on healthcare systems and patients’ quality of life. [15] A fundamental clinical challenge is that early-stage CKD frequently lacks overt symptoms, which impedes timely identification. Once clinical signs become apparent, renal injury has typically progressed to an irreversible stage. [16] Therefore, elucidating the underlying pathophysiological mechanisms and identifying high-risk individuals at an early stage are critical for preventing disease progression. [17] Growing evidence has underscored the pivotal contributions of chronic inflammation and nutritional disturbances to CKD pathogenesis. Chronic inflammation not only accelerates renal injury but also contributes to the development of protein-energy wasting in CKD patients, [18] forming an “inflammation–malnutrition” vicious cycle that further exacerbates renal function decline and heightens the risk of associated complications. [19] The HALP score, an integrated composite metric reflecting both inflammatory and nutritional status, has been linked to a variety of conditions, including overactive bladder syndrome, [20] coronary heart disease, [21] and psoriasis. [22] Specifically in the renal context, reduced HALP scores correlate with advanced tubular atrophy and interstitial fibrosis in IgAN. [23] However, prior studies have largely focused on specific renal disease subtypes, such as DN, IgAN, lupus nephritis, and dialysis populations, with relatively few investigations focusing on the general adult population using large nationally representative datasets. In the present NHANES analysis, we observed a robust inverse association between the HALP score and CKD risk, and notably identified a non-linear threshold effect that warrants further discussion.

The HALP score threshold identified in our analysis (52.43) is exploratory and requires external validation before clinical use. If validated, this cutoff could help clinicians identify patients who may benefit from nutritional intervention and regular inflammatory monitoring. Notably, similar non-linear, threshold-type associations have been reported for HALP in other disease contexts. Ding et al. [24] reported an L-shaped relationship between the HALP score and diabetic retinopathy, with risk declining sharply until plateauing above a threshold of 42.9. Similarly, a threshold of 44.98 was reported for diabetic foot ulcer risk [25]. These consistent threshold patterns across different diseases suggest that the HALP score may reflect common underlying pathophysiological mechanisms.

The differential associations of the HALP score with albuminuria and low-eGFR are biologically plausible and warrant further consideration. Albuminuria reflects early glomerular filtration barrier injury, whereas reduced eGFR typically manifests in later disease stages and indicates established structural damage. [26] The persistent inverse association with albuminuria after full adjustment, contrasted with the attenuated signal for low-eGFR, suggests that the HALP score may be particularly sensitive to early renal injury and therefore may help identify individuals with elevated CKD risk. Subgroup analyses further confirmed that the HALP score is independently associated with CKD across diverse populations, supporting its role as a complementary indicator to refine current risk evaluation models, particularly for high-risk individuals with hypertension, diabetes [27], or advanced age. [28]

The superior discriminative ability of the HALP score compared with PNI, PLR, LMR, and SII may reflect its integration of multiple potentially CKD-related pathophysiological pathways. Based on existing literature, decreased hemoglobin has been associated with renal hypoxia and activation of the sympathetic and renin-angiotensin-aldosterone systems, potentially aggravating kidney injury. [29] Second, serum albumin exhibits antioxidant, anti-inflammatory, and antiplatelet aggregation properties, which may contribute to renal protection. [30] Third, lymphocytes and their mediators are thought to help mitigate renal inflammation and fibrosis, and lymphopenia has been linked to immune dysregulation in CKD. [31] Fourth, activated platelets may promote CKD progression through microthrombosis formation, release of inflammatory mediators, and profibrotic signaling. [32] Collectively, the HALP score may integrate these mechanisms, potentially offering a more comprehensive assessment of renal injury risk than single-axis indices.

To validate the clinical findings, we established a 5/6 nephrectomy rat model. CKD rats exhibited typical renal dysfunction and characteristic pathological damage compared with controls, confirming successful model establishment. [33] Importantly, CKD rats showed a significantly decreased HALP score, accompanied by reduced Hb, ALB, and LYM levels and elevated PLT counts, consistent with concurrent inflammatory activation and nutritional deterioration that may be associated with renal injury initiation and progression. [34] Pearson correlation analysis further revealed significant negative correlations between the HALP score and Scr, BUN, and UACR, with UACR showing the strongest correlation, highly consistent with our clinical findings and reinforcing the utility of the HALP score for CKD prevention and screening. [35]

Building on the above findings, several strengths of this study deserve further emphasis. First, this work systematically evaluates the HALP-CKD relationship in a large, nationally representative general population, extending prior work that has been largely confined to specific renal disease subtypes (e.g., DN, IgAN, lupus nephritis, dialysis). Second, this exploratory threshold, once externally validated, could support large-scale screening among high-risk populations, including those with hypertension, diabetes or advanced age. Third, ROC comparisons demonstrated that the HALP score outperformed several established inflammation-nutrition indices (PNI, SII, LMR, PLR) for identifying CKD and albuminuria, supporting its added clinical value. Most importantly, the inclusion of a 5/6 nephrectomy rat model enhances the translational relevance of our findings. In CKD rats, the HALP score was reduced and showed a strong negative correlation with renal injury markers, particularly the UACR. These results provide experimental validation for the clinical findings and further support the association between the HALP score and CKD. This combination of population-based and animal evidence, which has been rarely reported in previous HALP related studies, strengthens both the credibility and biological plausibility of our conclusions.

Several limitations of this study should be acknowledged. First, the cross-sectional observational design of the NHANES dataset precludes causal inference regarding the association between the HALP score and CKD. Second, despite comprehensive adjustment for known confounders, residual confounding by unmeasured factors cannot be fully eliminated. Third, the proposed links between the HALP score and CKD—via inflammation, malnutrition, and platelet activation—remain speculative, because relevant mechanistic biomarkers and molecular pathways were not examined in this study. These hypotheses require validation in future experiments. Finally, although the animal data support the clinical findings, causality remains unestablished. Longitudinal studies and interventional trials targeting nutritional and inflammatory pathways are needed to determine whether modulating the HALP score can reduce CKD incidence or slow progression.

## 5 Conclusion

Our integrated population and animal data support the HALP score as a simple and clinically informative indicator associated with CKD prevalence. Further longitudinal and interventional studies are warranted to establish its predictive utility and to elucidate the underlying mechanisms.

## Data availability statement

Publicly accessible datasets from https://www.cdc.gov/nchs/nhanes were used in this study.

## Ethics statement

Adhering to the ethical principles outlined in the Declaration of Helsinki, this study received approval from the NCHS Ethics Review Board. All participants provided signed informed consent prior to enrollment. Approval for the animal study protocol was granted by the Animal Ethics Committee at Hebei Provincial Hospital of Traditional Chinese Medicine. (Approval No.: IACUC-HPHCM-2026015).

## Author contributions

**Conceptualization:** Zhiqiang Chen, Xueqin Zhang

**Data curation:** Wenfeng Ye, Yiman Wang

**Formal analysis:** Wenyu Zhang, Yashi Wang

**Investigation:** Wenfeng Ye, Yiman Wang

**Methodology:** Xuejing Chen, Bingwu Zhao

**Project administration:** Zhiqiang Chen, Xueqin Zhang

**Resources:** Zhiqiang Chen, Xueqin Zhang

**Supervision:** Zhiqiang Chen, Xueqin Zhang

**Visualization:** Bingwu Zhao, Xuejing Chen

**Writing – original draft:** Wenyu Zhang, Yashi Wang

**Writing – review & editing:** Wenyu Zhang, Yashi Wang

## Declaration of Non-Duplicate Publication

The authors declare that the work is original and not submitted to any other journal.

